# Differentiating standard geometry and hyperangulated video laryngoscope blades by handle-blade angle, blade curvature, and camera field of view

**DOI:** 10.64898/2026.10.22.26366035

**Authors:** Brian E Driver, Stacy A Trent, Stephanie C DeMasi, Aaron E Robinson, Matthew E Prekker

## Abstract

**Background:** Video laryngoscopes are classically grouped into standard geometry (Macintosh-style) and hyperangulated classes. However, this distinction rests on the original Glidescope’s historically cited 60° angle rather than any recognized definition of a blade’s “angle.” Different methods of measuring angulation of a blade yield angles that differ by 30° or more, and blades are classified by convention rather than by measurement. We developed standardized geometric measurements and determined which best discriminate between the two blade types.

**Methods:** In a bench study of 23 commercially produced adult laryngoscope blades, we measured (i) blade and camera angles relative to the handle, (ii) intrinsic blade curvature from heel to tip, and (iii) camera field of view. The primary analysis compared measurements from 11 standard geometry video laryngoscopes and 8 non-channeled, hyperangulated video laryngoscopes. We report medians and Hodges-Lehmann median differences with 95% confidence intervals.

**Results:** The handle-referenced blade tip angle did not distinguish the classes (median 40° standard geometry vs 48° hyperangulated; median difference 8°, 95% CI −5° to 16°), and the vertical video field of view did not differ (34° vs 37°; difference 3°, 95% CI −1° to 9°). The intrinsic blade curvature, measured without relation to the handle, differed between classes. Hyperangulated blades had a larger heel-to-tip deflection angle (80° vs 49°; difference 31°, 95% CI 21° to 40°) and a smaller average radius of curvature (70 vs 113 mm; difference −42 mm, 95% CI −57 to −28 mm) despite similar arc length (119 vs 120 mm).

**Conclusions:** Intrinsic blade curvature, rather than the field of view or often-cited angle between the handle and blade tip, best distinguished standard geometry from hyperangulated video laryngoscopes. Reporting shape-based geometry measurements would support consistent classification and comparison.

## INTRODUCTION

Video laryngoscopes are routinely divided into two classes: standard geometry (Macintosh-style) blades, which retain the shape of the traditional Macintosh direct laryngoscope,^1^ and hyperangulated blades, which are more sharply curved to visualize the larynx with less manipulation of upper airway soft tissues, precluding a direct line of sight. The prototypical hyperangulated video laryngoscope is the original Glidescope, described in its first clinical report as “similar to a Macintosh blade but the blade has a pronounced 60° angulation.”^2^ No measurement method or source accompanied this angulation claim, which has nonetheless been repeated for two decades as the defining characteristic of hyperangulated blades.^3–6^

More fundamentally, there is no accepted definition of the “angle” of a laryngoscope blade or how it should be measured. Published descriptions of individual devices have measured the angle between the handle and blade tip relative to a horizontal line drawn from the handle-mounting block (35° to 39° for a novel curved blade),^7^ the angle between the blade tip and the handle (58° for a Macintosh blade and 76° for the Dörges universal blade),^8^ separate “blade angulation” and “anterior view” angles for the C-MAC (18° to 30°, varying with blade size),^9^ and protractor photographs in which a Glidescope reaches 60° only when tipped backward on a tabletop, with the angle referenced to neither the handle nor the orientation of use (examples shown in Fig. S1 in the appendix).^10^ Others have instead referenced the blade’s view axis to the vertical axis of the handle, assigning non-channeled video laryngoscopes a line of sight of approximately 270° to 300° when the handle axis is taken as 0°.^11^ The international standard governing laryngoscopes, ISO 7376, specifies the handle-blade interface and illumination but describes blade shape only as “curved, straight, hybrid and others” and defines no angular measurement.^12^ Different measurement conventions yield angles differing by 30° or more in the same laryngoscope blade.

The distinction between standard geometry and hyperangulated blades is not merely semantic. Clinical trials, meta-analyses, and airway guidelines group devices by blade class, and reported outcomes, including glottic view, first-attempt success, and ease of tube delivery, differ by class. Intubation technique also differs: hyperangulated blades are typically paired with an endotracheal tube containing a rigid, angulated stylet,^5,11^ whereas standard geometry blades permit conventional Macintosh technique. Yet devices are assigned to these classes by convention and visual impression rather than by any quantitative criterion. Without standardized measurements, it is impossible to state where standard geometry ends and hyperangulation begins, whether devices within a class are geometrically comparable, or how newer “intermediate” blades should be classified.^4^ Furthermore, video laryngoscope classification using a single measurement may be too simplistic because devices may differ in curvature, where along the blade the curvature occurs, and in the camera’s field of view. These attributes all plausibly affect clinical performance but are rarely measured or reported.

Using a sample of contemporary standard geometry and hyperangulated laryngoscopes, we sought to develop and measure a standardized, reproducible set of geometric measurements of the angle of the blade tip relative to the handle, the intrinsic curvature of the laryngoscope blade, and the field of view of the video camera. Our goals were to quantify the geometric differences between the two classes and to determine which measurements best discriminate them.

## METHODS

### Study Design

We performed a bench study using a convenience sample of commercially produced adult laryngoscope blades, comprising standard geometry and hyperangulated video laryngoscopes as well as direct laryngoscope blades. Full detail is shown in Table S1 in the Supplementary Appendix.

The following 11 standard geometry video laryngoscope blades were tested: Ambu SureSight Mac 3, Ambu SureSight Mac 4, C-MAC Mac 3, C-MAC Mac 4, C-MAC S Mac 4 (single use), Glidescope Spectrum Mac S3, Glidescope Spectrum Mac S4, McGrath Mac 3, McGrath Mac 4, OneScope Pro-Direct Mac 4, and i-view.

The following 8 hyperangulated blades without a channel were tested: C-MAC D-blade, C-MAC S D-blade (plastic), Glidescope GS-1001-100 (from the original commercially marketed Glidescope platform, widely regarded as the first commercially available video laryngoscope), Glidescope GVL 3 Stat, Glidescope GVL 4 Stat, Glidescope Spectrum Hyperangle S3, Glidescope Spectrum Hyperangle S4, and McGrath X3.

We also measured blades included in additional analyses but not the main analysis, including two Macintosh direct laryngoscope blades (Macintosh direct laryngoscope size 3 and Macintosh direct laryngoscope size 4, both from Rusch Polaris), and two hyperangulated blades with a channel (KingVision and AirTraq).

The study did not involve human participants and did not require institutional review board approval.

### Methods of Measurement

Three domains of blade geometry were measured and are summarized below: (1) Angle of the blade tip and camera face relative to the handle (Figure 1); (2) Intrinsic curvature of the laryngoscope blade (Figure 2); and (3) Field of view of the video camera (Figure 3). Complete measurement protocols and the analytic definitions used are provided in the Supplementary Appendix.

**Figure 1:**
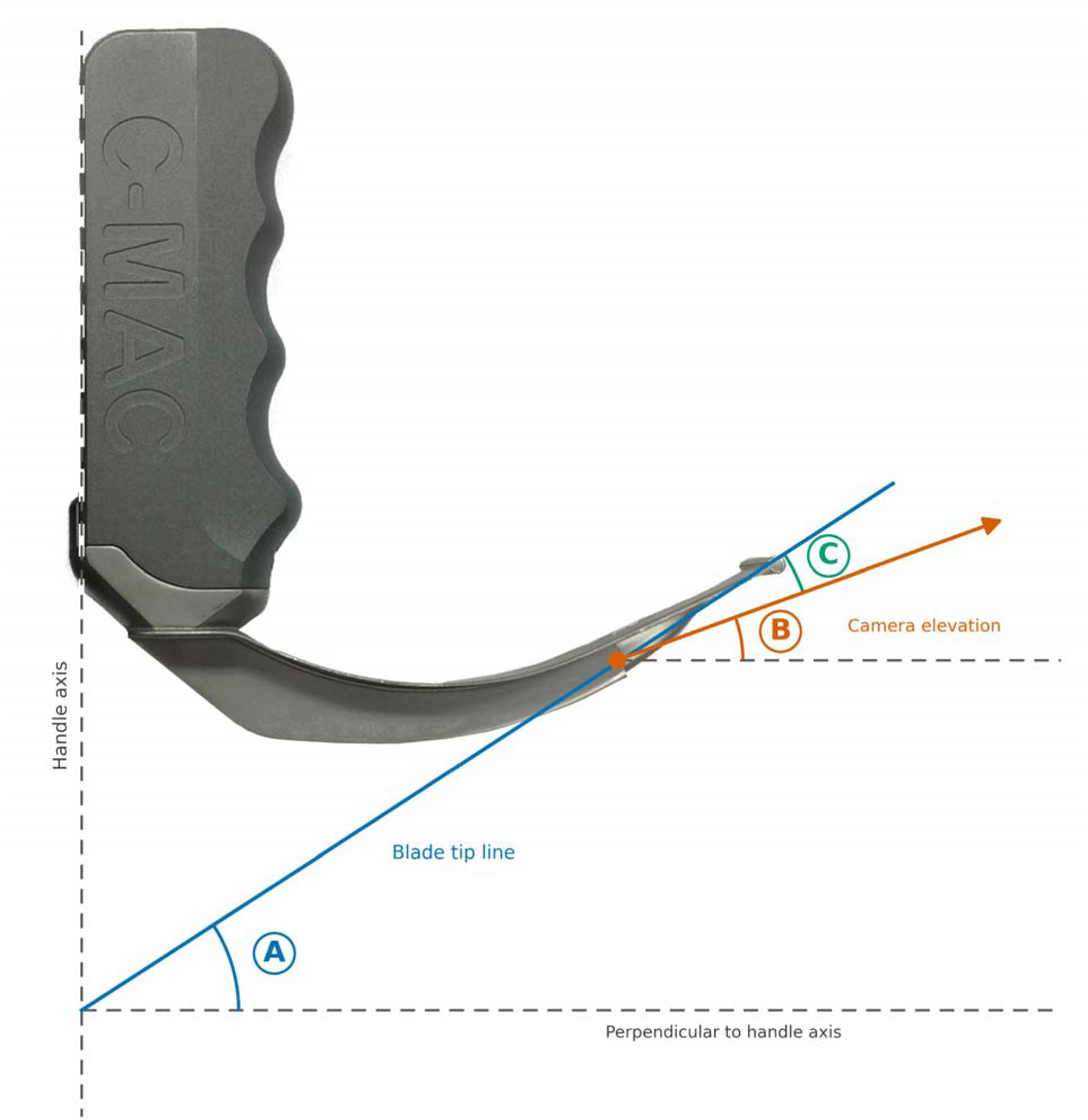
Blade and camera angles relative to the handle (Measures A–C) Angles of the blade tip and the camera face relative to the laryngoscope handle. The handle axis and its perpendicular are shown as dashed lines. A, Blade tip angle: the angle of the blade tip line above the perpendicular to the handle axis (computed as 90° minus the protractor angle between the handle axis and the blade tip line). B, camera face angle: the angle of the camera case above the same perpendicular, measured with the protractor arm perpendicular to the camera face. C, blade tip-camera angle (C = A − B). The photograph shows a C-MAC Mac 4 video laryngoscope; the dashed handle axis is drawn along the back of the handle; the drawn angles are illustrative.

**Figure 2:**
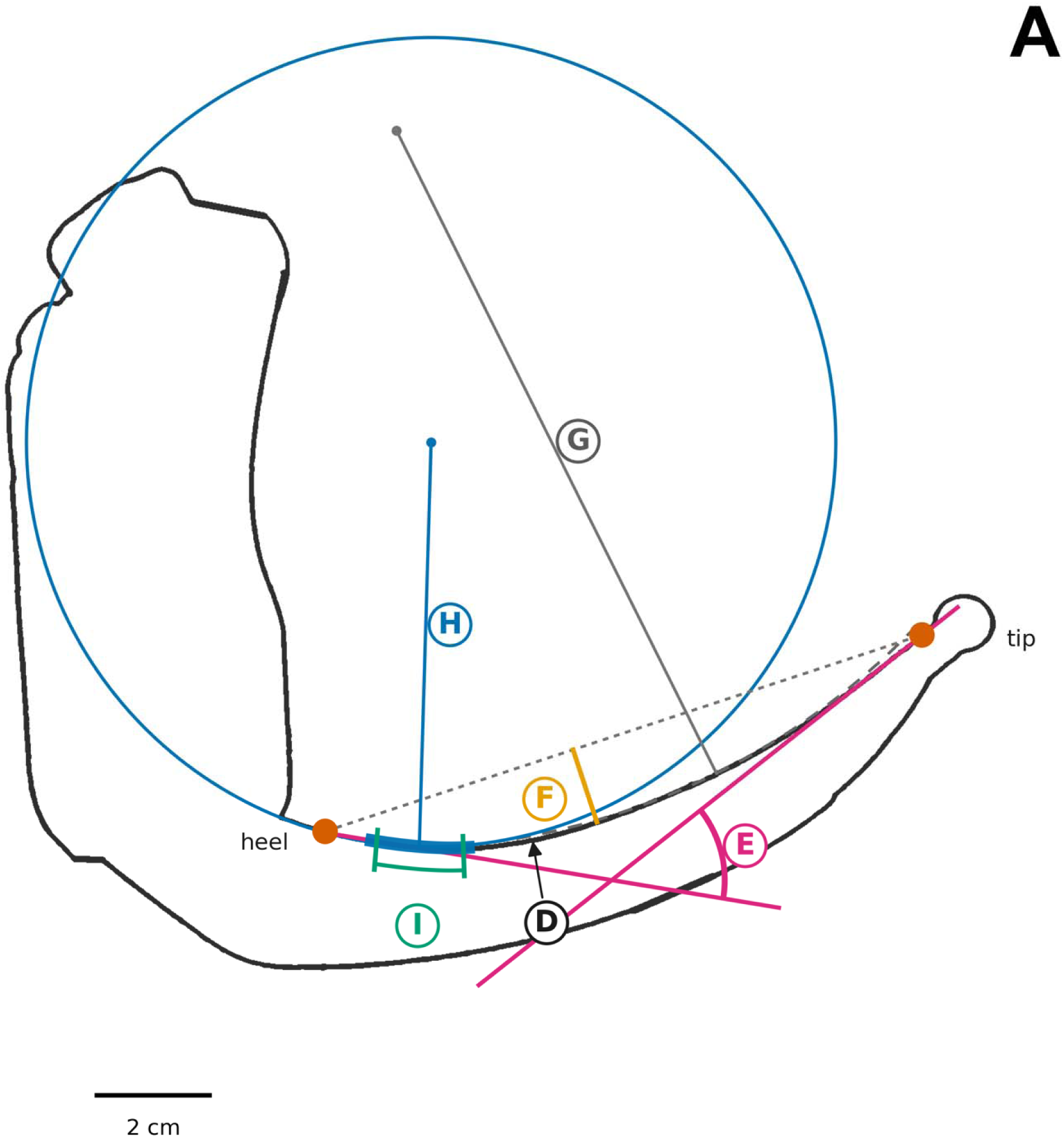
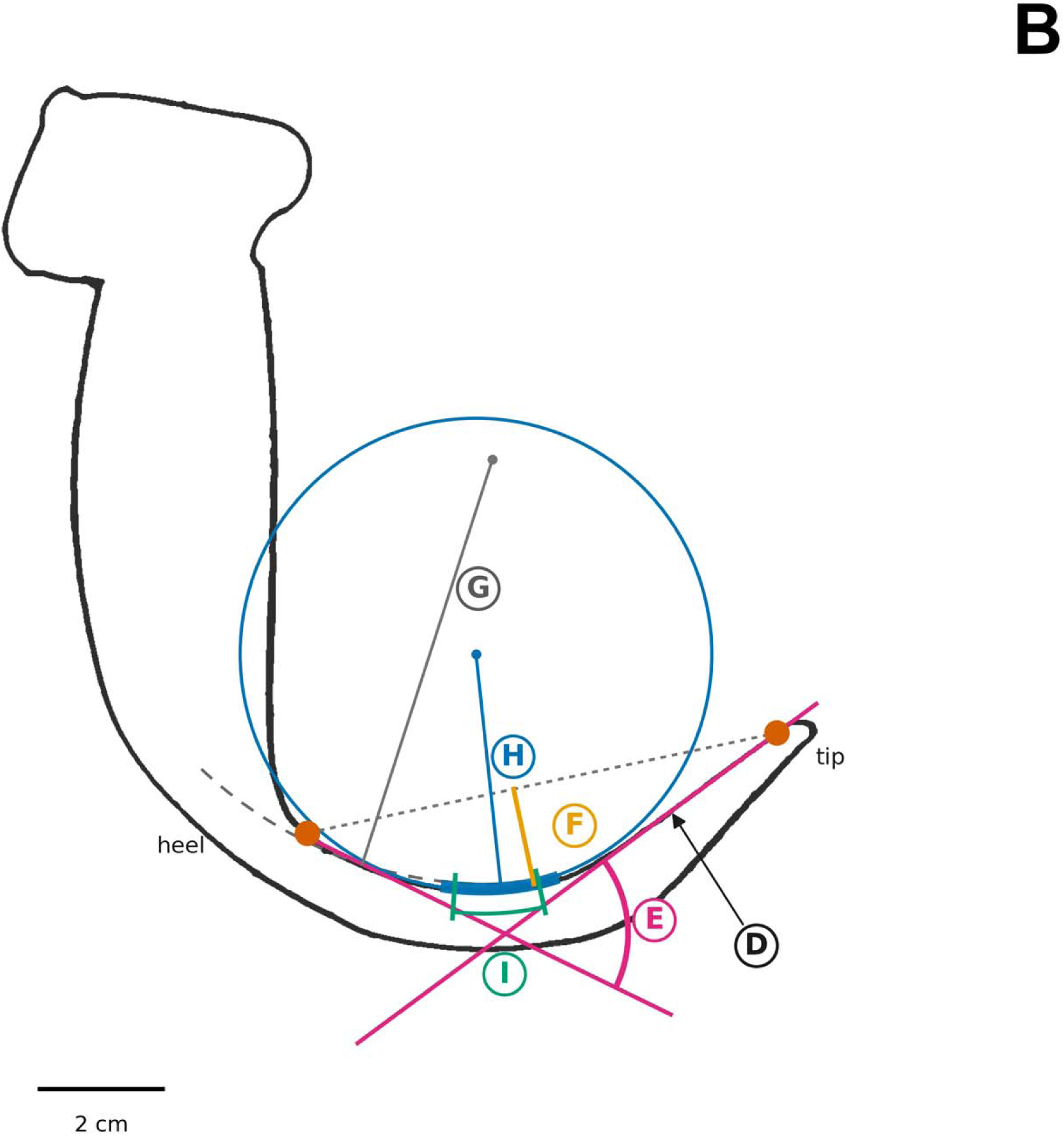
Intrinsic curvature of the blade, C-MAC Mac 4 and Glidescope Hyperangle S3 (Measures D–I) Intrinsic curvature measures of the traced concave (lingual) edge of the blade, shown in matching format on tracings of a C-MAC Mac 4 (standard geometry, Panel A) and a Glidescope Hyperangle S3 (hyperangulated, Panel B) so the two blade shapes can be compared directly. D, arc length of the traced concave edge between the dots marking the heel and tip. E, heel-to-tip deflection angle: the angle between the heel and tip segment tangents (intrinsic blade angulation). F, sagitta: the maximum perpendicular bow of the edge off the heel–tip chord (dotted); reported as the ratio sagitta ÷ chord length. G, average radius of curvature of the whole edge (dashed arc with radius line). H, minimum radius: the tightest bend sustained over a 2 cm window (window darkened on the edge; circle drawn at that radius). I, tight-bend zone: near and far edges of the region of tightest bending, measured from the tip.

**Figure 3:**
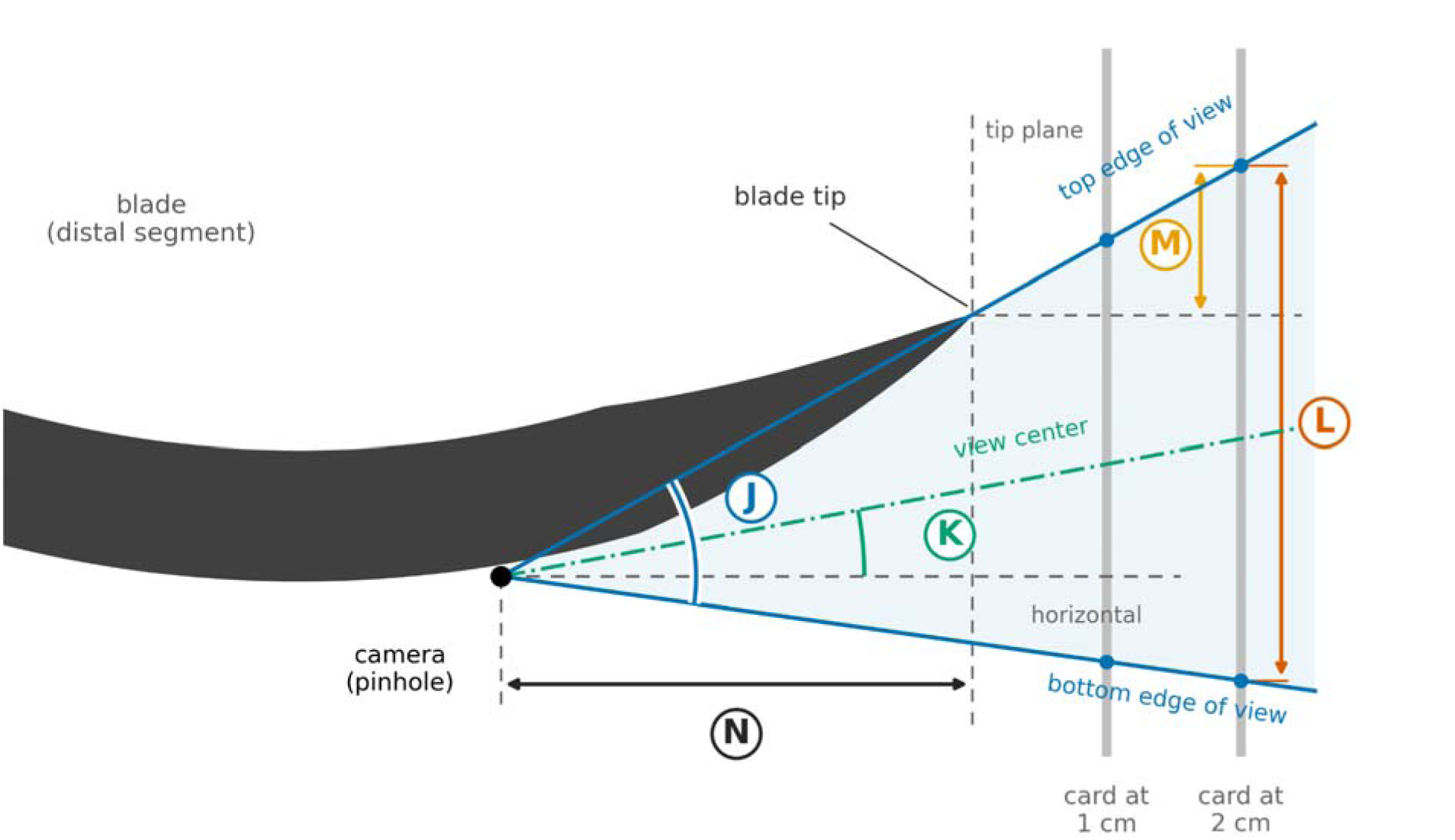
Field of view (Measures J–N) Video camera field-of-view geometry, side view (schematic; not to scale). The camera is treated as a pinhole, so each edge of the view is a straight ray. The handle was clamped in a vertical orientation, perpendicular to the dashed horizontal line. View edges were traced on vertical cards held 0, 1, and 2 cm beyond the blade tip (dots); edge-ray angles were reconstructed from the change between the 1 cm and 2 cm cards because at 0 cm the top edge of the view is occluded by the blade tip itself. J, vertical field of view: the angle between the top and bottom edges of the view. K, view center elevation: the angle of the view center above horizontal (positive = aimed upward). L, vertical span of the view on the card 2 cm beyond the tip. M, vertical distance from the top of the view down to blade-tip height, at 2 cm. N, camera setback behind the tip plane.

#### Angle of the blade tip and camera face relative to the handle

First, with the handle held vertical, we used a digital protractor to measure the raw angle between the vertical axis of the handle and the line parallel to the last centimeter of the blade tip. Subtracting this raw reading from 90° yields the blade tip angle—the elevation of the blade tip line above the horizontal plane (perpendicular to the vertical handle axis, measure A, Figure 1). Second, using the same technique, we measured the angle between the vertical axis of the handle and the face of the camera (with the protractor perpendicular to the camera face) and again subtracted it from 90° (measure B, Figure 1). Measures A and B were measured in triplicate and averaged. Third, we calculated the blade tip-camera angle (measure C, Figure 1) as measure A minus measure B (C = A − B).

#### Intrinsic curvature of the laryngoscope blade

To characterize blade curvature, we carefully traced the outline of each blade onto paper. The tracings were scanned, and curvature measures including arc length, heel-to-tip deflection angle (the intrinsic angulation of the blade, independent of the handle), sagitta, average and minimum radius of curvature, and the location of the tight-bending zone, were computed from the digitized concave edge using a custom Python program (measures D-I; Figure 2). The blade edge started in alignment with the front of the handle and ended where the distal blade meets the rounded blade tip. The tracing procedure, image calibration, and the definition of each curvature measure are described in the Supplementary Appendix.

#### Field of view of the video camera

To measure the camera field of view, each laryngoscope handle was clamped in a vertical orientation, and a flat paper card was taped to a vertical surface facing the camera. With the paper positioned at 0, 1, and 2 cm beyond the blade tip we measured the vertical span of the camera view. These spans were used to reconstruct the field-of-view geometry, including the vertical field of view, the elevation of the view center, and the camera setback behind the blade tip (measures J-N; Figure 3).

### Primary Analysis

The primary comparison was between standard geometry and hyperangulated video laryngoscope blades. Because the measurements were continuous and not assumed to be normally distributed, we summarized each characteristic as the median value and interquartile range within each geometry class. For each characteristic, we estimated the Hodges-Lehmann median difference between the hyperangulated and standard geometry blade groups with an accompanying 95% confidence interval (CI). In an additional analysis, we then repeated these comparisons after including direct Macintosh laryngoscopes in the standard geometry group and channeled hyperangulated blades in the hyperangulated group. Analyses were performed in Stata (StataCorp, College Station, TX, version 19).

## RESULTS

We evaluated 23 laryngoscope blades: 13 standard geometry blades (11 capable of video laryngoscopy, 2 direct laryngoscopy only) and 10 hyperangulated blades (all capable of video laryngoscopy; 8 non-channeled, 2 channeled) (see Table S1 for details). The primary analysis was a comparison of the 11 standard geometry video laryngoscopy blades versus 8 hyperangulated, non-channeled video laryngoscopy blades. The handle-referenced blade tip angle (Measure A)—the most commonly reported measurement—did not distinguish the two classes of blades (median 40° for standard geometry versus 48° for hyperangulated, median 8° difference, 95% CI −5° to 16°, Table 1 and Figure 4A). The camera face angle (Measure B) likewise did not differ, although it was far more variable among hyperangulated blades (Table 1, Figure 4B). The blade tip-camera angle (Measure C), however, differed between blade classes with hyperangulated class having a larger angle as compared to standard geometry class (30° vs 14°, median 14° difference, 95% CI 4° to 22°, Table 1).

**Figure 4:**
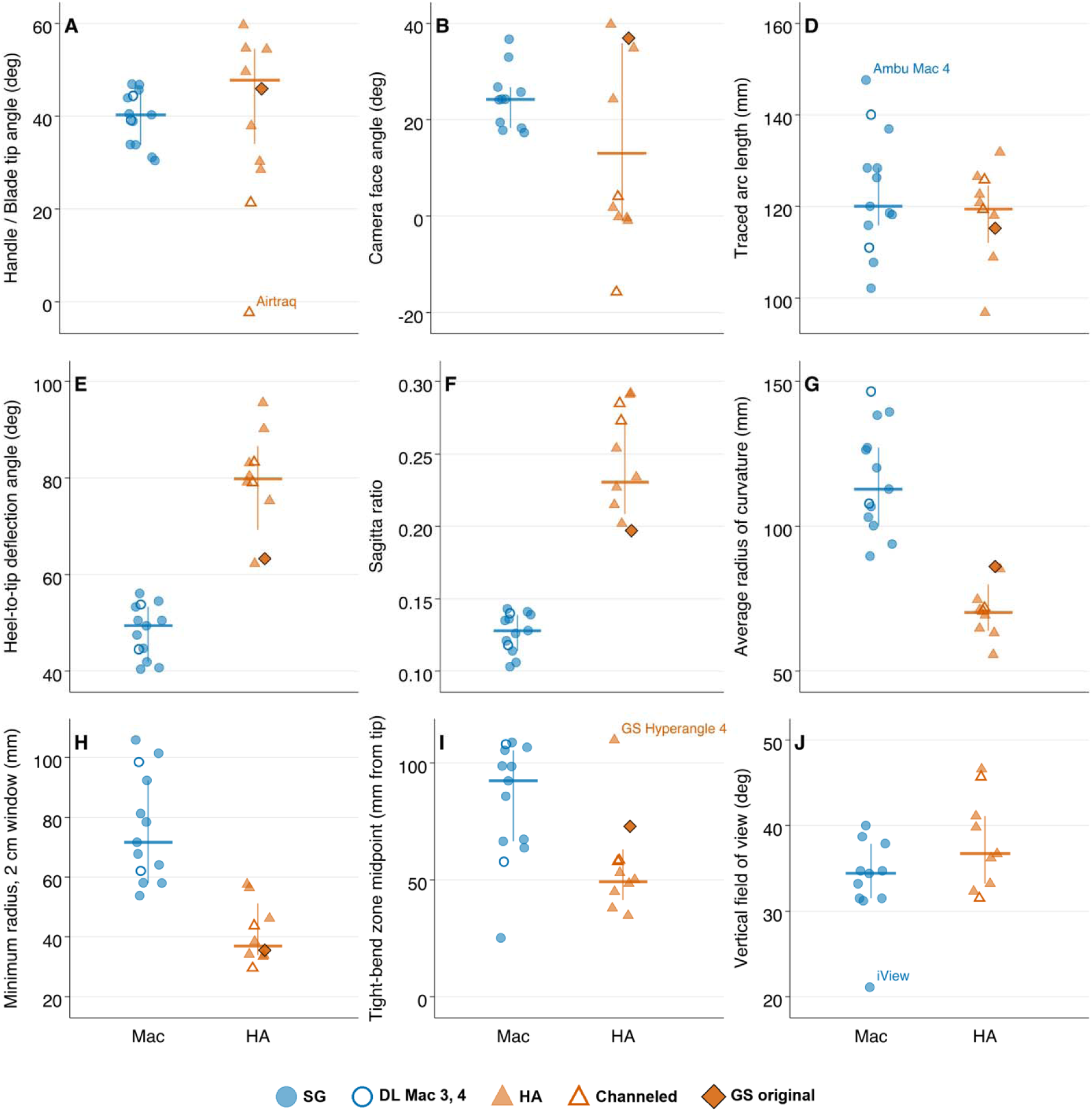
Handle-referenced angles, intrinsic curvature, and video camera field of view by blade categorization. Comparison of selected handle-referenced angles, intrinsic curvature, and video camera field of view measures between standard geometry (SG) and hyperangulated (HA) laryngoscope blades. Figure letters correspond to diagram location as shown in Table 1 and Figures 1-3. (A) Blade tip angle. (B) Camera face angle. (D) Traced arc length. (E) Heel-to-tip deflection angle (intrinsic blade angulation). (F) Sagitta ratio. (G) Average radius of curvature. (H) Minimum radius, 2 cm window. (I) Tight-bend zone midpoint. (J) Vertical field of view. Each point is one blade, jittered horizontally within its group. The horizontal crossbar and vertical whisker mark the group median and interquartile range (IQR: 25th-75th percentile). Marker key: filled blue circle = standard geometry blade; filled orange triangle = hyperangulated blade. Hollow blue circles denote the DL Mac 3 and DL Mac 4 blades. Hollow orange triangles denote channeled hyperangulated blades (Airtraq, KingVision). The filled orange diamond denotes the original Glidescope blade. For the standard geometry group, the median/IQR reflects only the video standard geometry blades. For the hyperangulated group, the median/IQR crossbar reflects only the non-channeled blades. Points labeled by blade name are outliers, with values beyond 1.5x the interquartile range from the group’s 25th or 75th percentile. Sample sizes vary slightly by panel because of missing measurements for individual blades. Overall n = 13 standard geometry, 10 hyperangulated, except: panel B (camera face angle) excludes DL Mac 3 and DL Mac 4, which have no camera; panel J (vertical field of view) excludes DL Mac 3, DL Mac 4, and the original Glidescope blade, for which field of view data were not available.

**Table 1:**
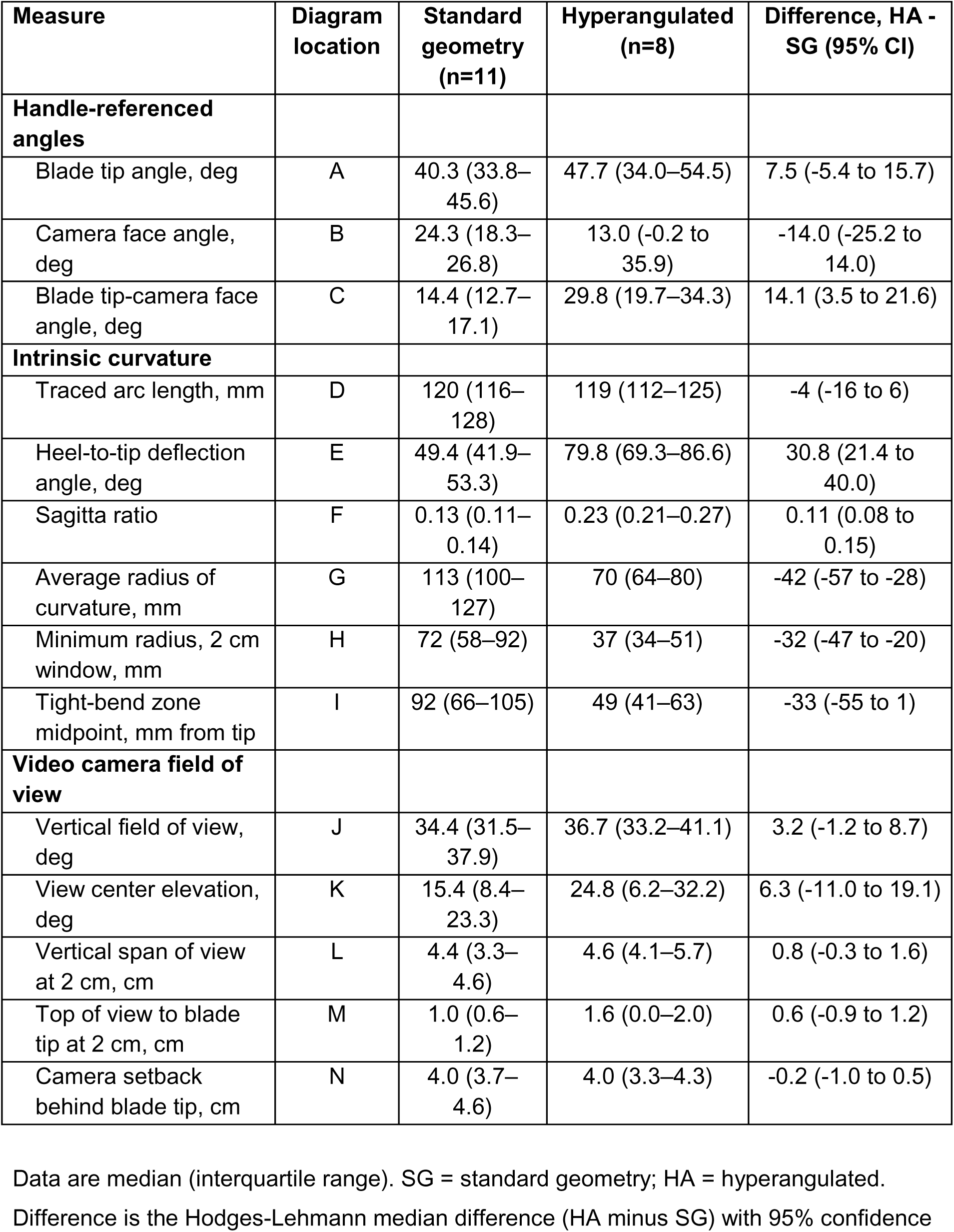
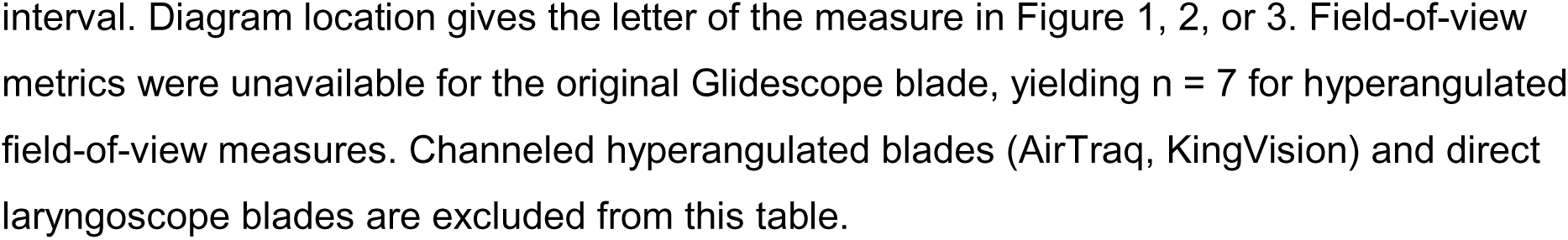
Handle-referenced angles, intrinsic curvature, and field-of-view characteristics of standard geometry versus hyperangulated laryngoscope blades.

| Measure | Diagram location | Standard geometry (n=11) | Hyperangulated (n=8) | Difference, HA - SG (95% CI) |
| --- | --- | --- | --- | --- |
| **Handle-referenced angles** |  |  |  |  |
| Blade tip angle, deg | A | 40.3 (33.8–45.6) | 47.7 (34.0–54.5) | 7.5 (-5.4 to 15.7) |
| Camera face angle, deg | B | 24.3 (18.3–26.8) | 13.0 (-0.2 to 35.9) | -14.0 (-25.2 to 14.0) |
| Blade tip-camera face angle, deg | C | 14.4 (12.7–17.1) | 29.8 (19.7–34.3) | 14.1 (3.5 to 21.6) |
| **Intrinsic curvature** |  |  |  |  |
| Traced arc length, mm | D | 120 (116–128) | 119 (112–125) | -4 (-16 to 6) |
| Heel-to-tip deflection angle, deg | E | 49.4 (41.9–53.3) | 79.8 (69.3–86.6) | 30.8 (21.4 to 40.0) |
| Sagitta ratio | F | 0.13 (0.11–0.14) | 0.23 (0.21–0.27) | 0.11 (0.08 to 0.15) |
| Average radius of curvature, mm | G | 113 (100–127) | 70 (64–80) | -42 (-57 to -28) |
| Minimum radius, 2 cm window, mm | H | 72 (58–92) | 37 (34–51) | -32 (-47 to -20) |
| Tight-bend zone midpoint, mm from tip | I | 92 (66–105) | 49 (41–63) | -33 (-55 to 1) |
| **Video camera field of view** |  |  |  |  |
| Vertical field of view, deg | J | 34.4 (31.5–37.9) | 36.7 (33.2–41.1) | 3.2 (-1.2 to 8.7) |
| View center elevation, deg | K | 15.4 (8.4–23.3) | 24.8 (6.2–32.2) | 6.3 (-11.0 to 19.1) |
| Vertical span of view at 2 cm, cm | L | 4.4 (3.3–4.6) | 4.6 (4.1–5.7) | 0.8 (-0.3 to 1.6) |
| Top of view to blade tip at 2 cm, cm | M | 1.0 (0.6–1.2) | 1.6 (0.0–2.0) | 0.6 (-0.9 to 1.2) |
| Camera setback behind blade tip, cm | N | 4.0 (3.7–4.6) | 4.0 (3.3–4.3) | -0.2 (-1.0 to 0.5) |

Data are median (interquartile range). SG = standard geometry; HA = hyperangulated.
Difference is the Hodges-Lehmann median difference (HA minus SG) with 95% confidence

The most consistent differences between the standard geometry and hyperangulated blade groups were observed in blade curvature (Measures E through I; Table 1, Figure 4E through 4I). The hyperangulated blades themselves, without relation to the handle, had a larger heel-to-tip deflection angle than standard geometry blades (median 80° versus 49°, Measure E) and were more tightly curved, with smaller average (70 versus 113 mm, Measure G) and minimum (37 versus 72 mm, Measure H) radii of curvature. Traced arc length was similar between classes (Measure D). Figure 5 shows an overlay of two blades, one in each class, that helps illustrate the actual differences between blade types.

**Figure 5:**
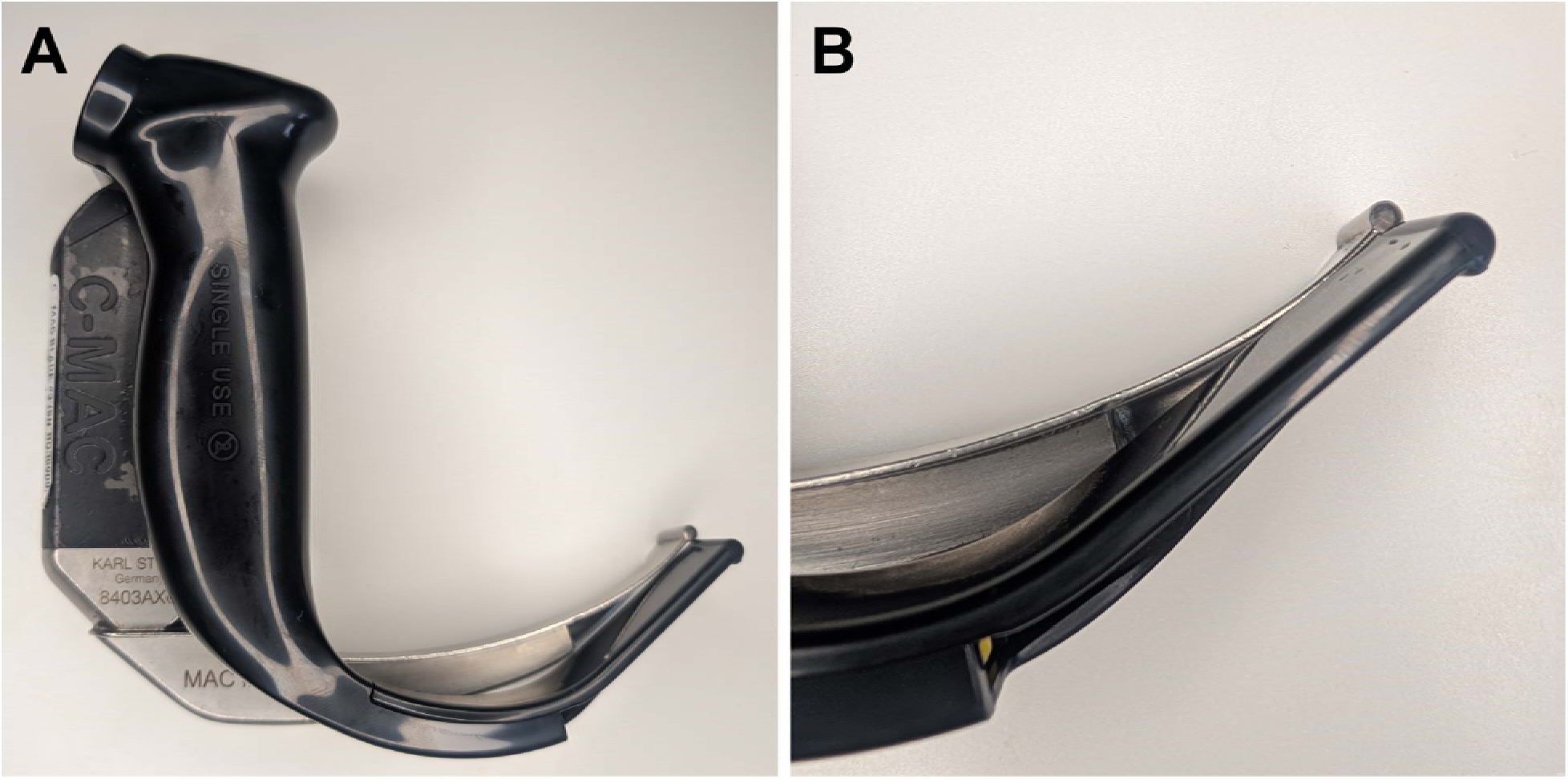
Standard geometry and hyperangulated video laryngoscope blade overlay. Glidescope Spectrum Hyperangle size 3 (black) overlaid on a C-MAC Macintosh size 3 (silver/black) with vertical handle axes aligned (A: full image; B: zoomed tip). While the handle-referenced blade tip angles (37.9° vs. 38.9°, Measure A in Fig. 1) and arc lengths (97 mm vs. 102 mm, Measure D in Fig. 2) are similar, the hyperangulated blade features a sharper heel-to-tip deflection angle (62° vs 40°, Measure E in Fig. 2) and a tighter radius of curvature (69 mm vs. 107 mm, Measure G in Fig. 2).

The original GlideScope blade (GS-1001-100), the device whose cited 60° angulation defined the hyperangulated class, had a handle-referenced blade tip angle of 46° (Measure A), an arc length of 115 mm (Measure D), a heel-to-tip deflection angle of 63° (Measure E), and an average radius of curvature of 86 mm (Measure G).

Camera field-of-view measures, including vertical field of view, view center elevation, and camera setback behind the blade tip, did not differ between the two classes, although the confidence intervals were wide (Measures J–N, Table 1, Figure 4J). The direction of the camera face was a poor guide to where the camera looked. Relative to the handle, the median camera face was aimed lower in hyperangulated blades (13° vs 24°, Measure B), whereas the median center of the video image was aimed higher (25° vs 15°, Measure K), although neither confidence interval excluded zero. Instead, the view center followed the blade tip, sitting a median of 25° below the blade tip line in both classes.

When the direct laryngoscopes were included with the standard geometry group and the channeled hyperangulated laryngoscopes (AirTraq and KingVision) included with the hyperangulated group, the spread of the hyperangulated angle measurements was wider but the curvature measurements were largely unchanged (Table S2).

## LIMITATIONS

This study has several limitations. First, it was designed to quantitatively compare laryngoscope blade geometries, not to link that geometry to clinical performance. The measurements we report do not necessarily correspond to the ease or difficulty of intubation at the bedside, which also depends on patient anatomy, operator technique, stylet use, and other factors that a bench study cannot capture. Relatedly, we do not know which specific blade features, if any, drive higher or lower intubation success, or whether the differences we measured are clinically meaningful. These findings should therefore be read as a geometric characterization of the blade classes rather than as evidence favoring any particular design.

Second, we compared only blades that fall clearly into the standard geometry or hyperangulated classes, as defined by manufacturers and clinical guidelines. We did not test intermediate or “hybrid” blades, whose geometry may lie between these groups. Third, the classification used to assign blades to a particular group was by convention rather than by an independent standard; the finding that curvature discriminates the classes therefore reflects, in part, that curvature approximates the intuitive basis on which blades are already sorted. Fourth, the sample was a convenience selection of commercially produced blades, with a single physical unit of each device, so it does not capture the full range of marketed laryngoscopes, and the small number of blades per class limits the precision of the estimated differences. Fifth, because we measured the camera face and the optical axis separately, we could not determine whether the difference between them reflects lens tilt, window geometry, or lens design. Finally, the measurements were made on isolated blades in a fixed bench orientation by a single investigator; although each angle was measured in triplicate and every curvature tracing was saved for reproducibility, some subjectivity in tracing and alignment remains and inter-rater reliability was not formally assessed. Field of view was derived from a simplified pinhole model on flat cards and does not reflect in vivo optics, image processing, the fullness of the displayed image, or the horizontal field of view.

## DISCUSSION

In this bench study of a sample of commonly used contemporary adult laryngoscopes, the geometric feature most often invoked to define hyperangulation was not the feature that distinguished hyperangulated from standard geometry blades. The blade tip and camera angles measured relative to the handle, which are the measurements often cited as the canonical *60°* of the original Glidescope, overlapped substantially between the two classes. The video field of view likewise did not differ. Instead, the intrinsic shape of the blade (measures E through I), measured without reference to the handle, was more consistently different between classes. Hyperangulated blades were more sharply deflected from the proximal to distal blade, more deeply bowed, and more tightly curved. The two classes had a similar blade arc length, indicating that hyperangulated blades fold a similar span of blade into a tighter curve (Fig. 5).

The handle-referenced blade tip angle historically used to name this class of devices^2,4,10,11^ turns out to be among the poorest at identifying its members, including the original GlideScope blade. None of its handle-referenced angles approached the 60° cited in its first clinical report; its blade tip angle was 46°, within the range of contemporary standard geometry blades. The only measure that came close was the heel-to-tip deflection angle of 63°, which suggests that the original description, whatever its method, captured the intrinsic bend of the blade rather than its relation to the handle.

Although some hyperangulated blades had handle-blade tip angles near 60°, this measure varied substantially within the class and as a standalone measure did not reliably differentiate them from standard geometry blades. Thus, reporting a single handle-referenced angle, as most prior descriptions and the original Glidescope report have done, conflates the handle-blade tip angle with intrinsic blade shape and does not support a consistent definition.

The shape differences we measured are likely the physical basis for the clinical behavior of these devices, familiar to experienced clinicians. A blade that curves tightly and looks around the corner precludes direct laryngoscopy, thereby mandating an indirect video or optical view. Thus, a functional definition of a hyperangulated blade may be the inability for the blade to be used for direct laryngoscopy. This is intuitive and clinically meaningful but cannot be applied before a blade is used and does not permit gradation. With a tight blade curve, laryngeal exposure is usually the easy step and tube delivery the hard one because the tube must be maneuvered around the blade.^2,11^ The hypothesized improved visualization of the vocal cords may explain preferential use of the hyperangulated blade for patients with a suspected difficult airway in some centers. However, the superiority of the hyperangulated blade versus standard geometry blade on grade of view and successful intubation on the first attempt overall, and among patients with an anticipated difficult airway, has not yet been proven in a randomized trial in patients outside of the operating room.

Trials,^13^ meta-analyses,^14,15^ and airway guidelines^16,17^ currently pool devices under the labels “Macintosh” (synonymous with “standard geometry”) and “hyperangulated,” treating each class as homogeneous and defined. However, the present data show meaningful within-class variation. Standardized reporting of key blade geometry measurements, ideally provided by manufacturers, would allow clinicians to better understand and differentiate laryngoscope blades and investigators to stratify or adjust for actual blade shape in comparative effectiveness research. Because blade geometry exists on a continuous spectrum spanning models and sizes, perhaps blade curvature behaves as a “dose.” For example, tighter curvature may facilitate better laryngeal views but greater difficulty of tube delivery. This would also help place “intermediate” blades on a continuum rather than in an undefined third category, as they are now. Standardization would also help reconcile heterogeneity across studies: existing data have found similar first-attempt success or conflicting results for standard geometry and hyperangulated video laryngoscopy after adjustment,^14,18,19^ a comparison that is difficult to interpret when the compared categories are themselves inconsistently constituted.

The blade tip-camera angle (Measure C) and the optical measurements of the camera view lead to different conclusions. Measure C, taken from the camera face with a protractor, was wider in hyperangulated blades (30° vs 14°), which suggests that their cameras look farther below the blade tip. But when measured from the video image itself, the center of the view sat a median of 25° below the blade tip line in both classes. The camera face angle reflects the orientation of the camera housing, whereas the field of view reflects the optical axis of the lens behind it. Thus, the orientation of the camera face is a poor proxy for where the camera looks, and device descriptions should report the optical view axis rather than the camera face angle.

In conclusion, in a sample of commonly used laryngoscopes, hyperangulated and standard geometry blades were best differentiated by the intrinsic shape of the blade, without reference to the handle, rather than the often-cited blade tip angle or the video field of view. Defining and reporting laryngoscope geometry by these shape measures, rather than by a single handle-referenced angle or a class label, would allow a more consistent and likely meaningful comparison of these devices in clinical practice, training, and research.

## Data Availability

All data produced in the present study are available upon reasonable request to the authors

## Author contributions

All authors conceived of the study. BD performed the measurements and data analysis and wrote the first manuscript draft. All authors contributed substantially to its revision.

## Acknowledgements

None

## Declaration of generative artificial intelligence (AI) in scientific writing

During the preparation of this work the authors used Claude in order to improve language and readability. After using this tool, the authors reviewed and edited the content as needed and take full responsibility for the content of the publication.

## SUPPLEMENTARY APPENDIX

**Table S1:**
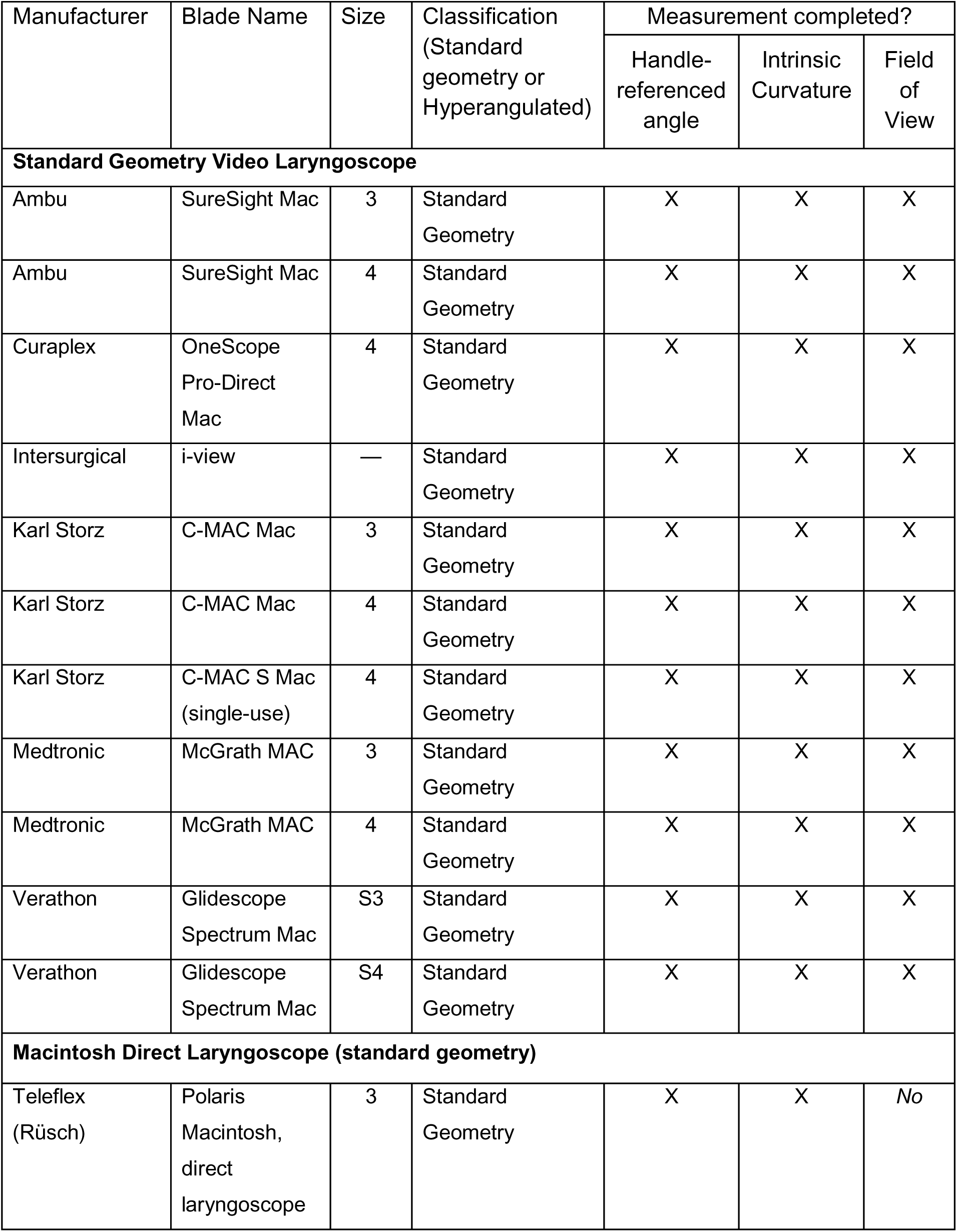
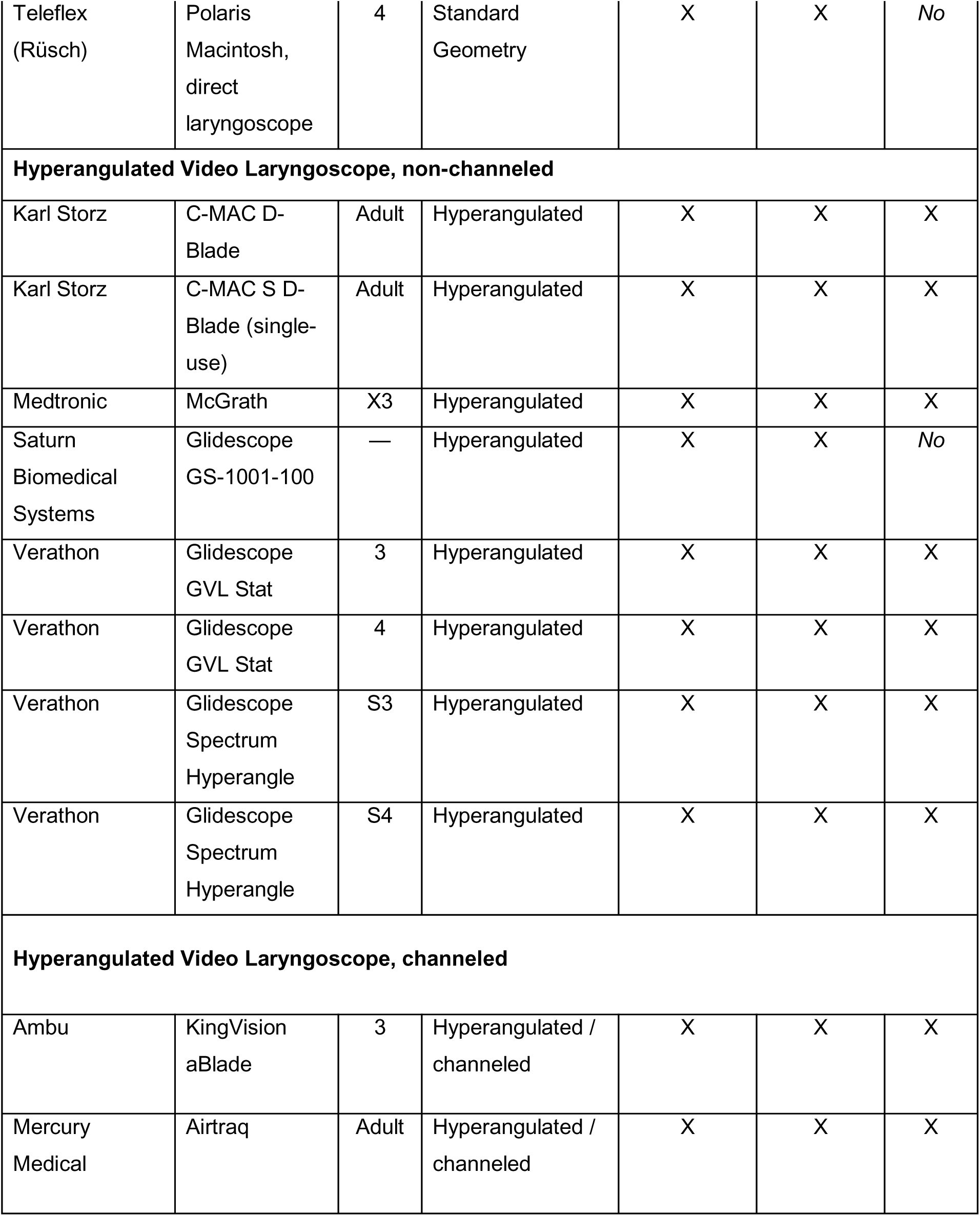
List of Laryngoscopes Tested.

**Table S2:**
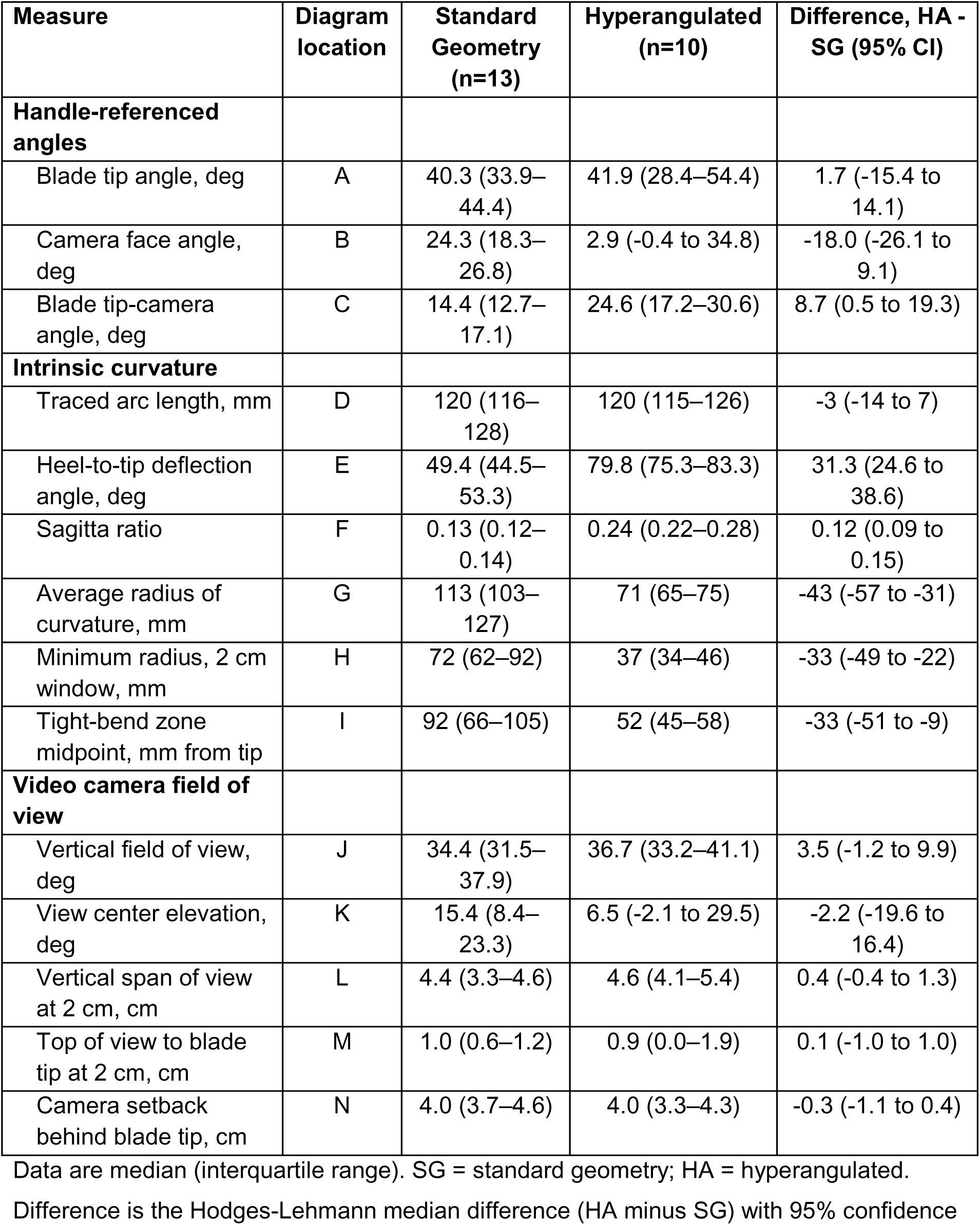
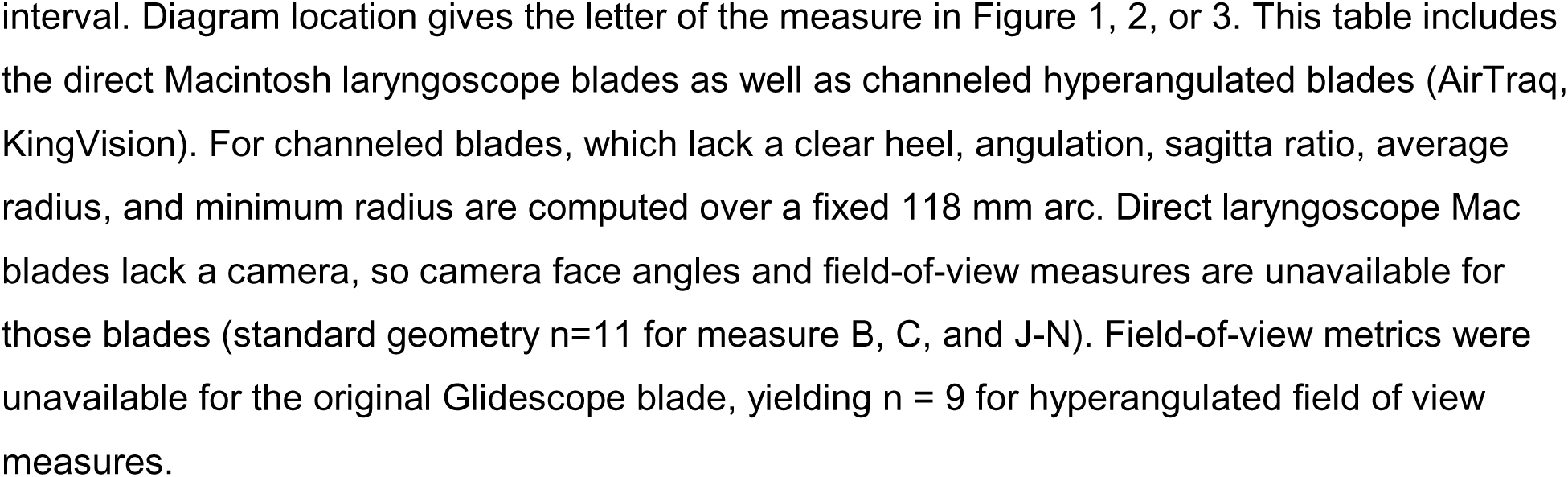
Handle-referenced angles, intrinsic curvature, and field-of-view characteristics of standard geometry versus hyperangulated laryngoscope blades (including channeled and direct blades)

| Measure | Diagram location | Standard Geometry (n=13) | Hyperangulated (n=10) | Difference, HA - SG (95% CI) |
| --- | --- | --- | --- | --- |
| **Handle-referenced angles** |  |  |  |  |
| Blade tip angle, deg | A | 40.3 (33.9–44.4) | 41.9 (28.4–54.4) | 1.7 (-15.4 to 14.1) |
| Camera face angle, deg | B | 24.3 (18.3–26.8) | 2.9 (-0.4 to 34.8) | -18.0 (-26.1 to 9.1) |
| Blade tip-camera angle, deg | C | 14.4 (12.7–17.1) | 24.6 (17.2–30.6) | 8.7 (0.5 to 19.3) |
| **Intrinsic curvature** |  |  |  |  |
| Traced arc length, mm | D | 120 (116–128) | 120 (115–126) | -3 (-14 to 7) |
| Heel-to-tip deflection angle, deg | E | 49.4 (44.5–53.3) | 79.8 (75.3–83.3) | 31.3 (24.6 to 38.6) |
| Sagitta ratio | F | 0.13 (0.12–0.14) | 0.24 (0.22–0.28) | 0.12 (0.09 to 0.15) |
| Average radius of curvature, mm | G | 113 (103–127) | 71 (65–75) | -43 (-57 to -31) |
| Minimum radius, 2 cm window, mm | H | 72 (62–92) | 37 (34–46) | -33 (-49 to -22) |
| Tight-bend zone midpoint, mm from tip | I | 92 (66–105) | 52 (45–58) | -33 (-51 to -9) |
| **Video camera field of view** |  |  |  |  |
| Vertical field of view, deg | J | 34.4 (31.5–37.9) | 36.7 (33.2–41.1) | 3.5 (-1.2 to 9.9) |
| View center elevation, deg | K | 15.4 (8.4–23.3) | 6.5 (-2.1 to 29.5) | -2.2 (-19.6 to 16.4) |
| Vertical span of view at 2 cm, cm | L | 4.4 (3.3–4.6) | 4.6 (4.1–5.4) | 0.4 (-0.4 to 1.3) |
| Top of view to blade tip at 2 cm, cm | M | 1.0 (0.6–1.2) | 0.9 (0.0–1.9) | 0.1 (-1.0 to 1.0) |
| Camera setback behind blade tip, cm | N | 4.0 (3.7–4.6) | 4.0 (3.3–4.3) | -0.3 (-1.1 to 0.4) |

Data are median (interquartile range). SG = standard geometry; HA = hyperangulated.
Difference is the Hodges-Lehmann median difference (HA minus SG) with 95% confidence

**Figure S1:**
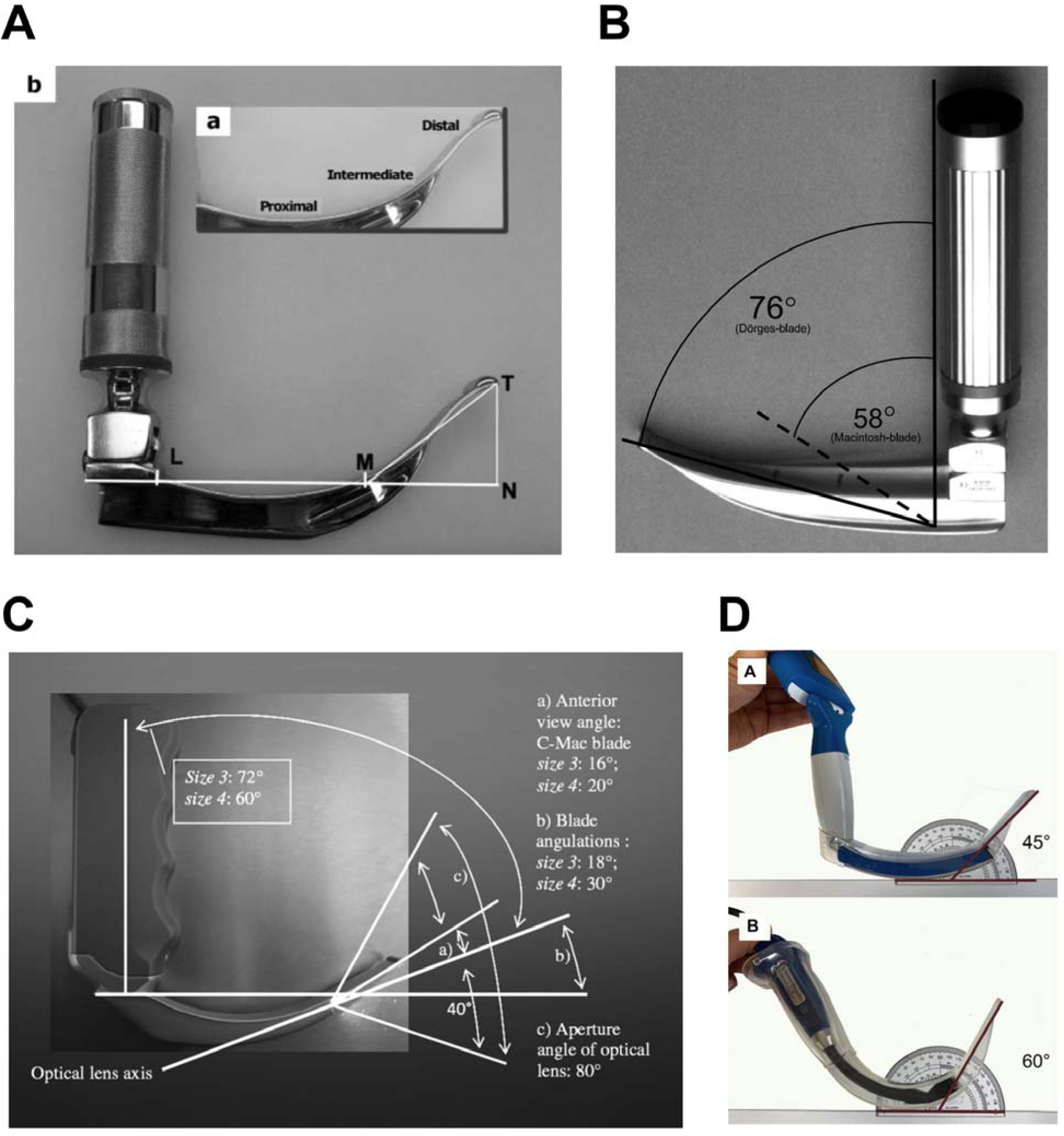
Examples of prior blade angle measurements. Published descriptions of individual devices have measured the angle between the handle and blade tip relative to a horizontal line drawn from the handle-mounting block (35° to 39° for a novel curved blade) (Panel A),^7^ the angle between the blade tip and the handle (58° for a Macintosh blade and 76° for the Dörges universal blade) (Panel B),^8^ separate “blade angulation” and “anterior view” angles for the C-MAC (18° to 30°, varying with blade size) (Panel C),^9^ and protractor photographs in which a Glidescope reaches 60° only when tipped backward on a tabletop, with the angle referenced to neither the handle nor the orientation of use (Panel D).^10^

## SUPPLEMENTARY METHODS

### Additional detail on measurements of the blade, handle, blade curvature, and field of view

#### Angle of the blade tip and camera relative to the handle

Angles were measured with a digital angle finder (GemRed 82305, 200 mm; Guilin GemRed Sensor Technology Co., Ltd), a stainless-steel ruler-and-protractor combination with a rated accuracy of ±0.3° and a locking function to hold a reading. With the laryngoscope handle held in a vertical orientation, one arm of the angle finder was aligned with the vertical axis of the handle and the other parallel with the last centimeter of the blade tip; the reading is the angle between the handle axis and the blade tip line. Subtracting this reading from 90° gives the handle-referenced blade tip angle relative to the horizontal plane (Measure A). The same procedure was used to measure the angle between the handle vertical axis and the camera face (Measure B). For this measure, the protractor arm was placed perpendicular to the face of the camera. Each angle was measured three times and the readings were averaged. The blade tip-camera angle (Measure C) was not measured directly but calculated based on the other two measured angles.

#### Intrinsic curvature of the laryngoscope blade

Each blade was traced by hand onto paper next to a 5 cm scale bar, and the tracing was scanned. On the scanned image, the investigator marked the two ends of the scale bar to calibrate the image in millimeters and then marked the heel and tip ends of the concave (lingual) edge of the blade. A custom Python program then followed the inked line between these two points automatically. If the program strayed from the blade edge, the investigator added guide points to correct the path. The traced edge was smoothed with a Gaussian filter with a standard deviation of 6 mm to remove small irregularities from hand tracing. Every smoothed tracing passed a quality check confirming that the edge curved in a single direction, defined as a total amount of turning along the edge no more than 15% greater than the net turning from heel to tip. The following measures were calculated from the smoothed edge:

D. Arc length is the length of the traced edge from heel to tip.
E. Heel-to-tip deflection angle is the angle between the direction of the first 20 mm of the edge at the heel and the last 20 mm at the tip. It reflects how far the blade turns from heel to tip, independent of the handle.
F. Sagitta ratio is the greatest distance between the edge and a straight line drawn from heel to tip, divided by the length of that line. It describes how deeply the blade bows and closely tracks the deflection angle, but it does not depend on the direction of the blade end.
G. Average radius of curvature is the radius of a circular arc with the same length and the same total turn as the blade edge, calculated as the arc length divided by the total turn from heel to tip. A smaller radius indicates a more tightly curved blade.
H. Minimum radius of curvature is the tightest bend sustained over any 2 cm stretch of the edge. A 2 cm window was moved along the edge, the radius within each window position was calculated in the same way as the average radius, and the smallest value was recorded.
I. Tight-bend zone location is the midpoint between the nearest and farthest points from the tip at which the 2 cm window radius was within 10% of the minimum radius.

For the channeled blades (AirTraq and KingVision), which lack a clearly defined heel, the deflection angle, sagitta ratio, and average and minimum radius were calculated over the distal 118 mm of the edge, measured back from the tip, so that the arbitrary choice of starting point did not affect these measures.

The calibration points, endpoints, guide points, and traced path were saved for each blade, along with an image of the trace overlaid on the scan, so that every measurement can be regenerated without retracing.

#### Field of view of the video camera

Each laryngoscope was clamped in a vise with the handle in a vertical orientation, and a flat paper card was taped to a vertical surface facing the camera. At distances of 0, 1, and 2 cm beyond the blade tip we marked the vertical span of the camera view visible on the card and the vertical distance from the top edge of the view down to the height of the blade tip.

The camera was treated as a pinhole, so each edge of the view travels in a straight line from the camera and rises or falls by a fixed amount for each centimeter of distance. The change in an edge’s height between the 1 cm and 2 cm cards therefore gives its angle, calculated as the arctangent of that change. All angles were measured relative to horizontal, defined as perpendicular to the vertical handle, with positive values aimed upward. The following measures were calculated:

J. Vertical field of view is the angle between the top and bottom edges of the view.
K. View center elevation is the angle of the center of the view, halfway between the top and bottom edges.
L. Vertical span at 2 cm is the distance between the top and bottom edges of the view on the 2 cm card.
M. Top of view to blade tip at 2 cm is the vertical distance on the 2 cm card from the top edge of the view down to the height of the blade tip.
N. Camera setback is the distance behind the blade tip at which the top and bottom edges of the view, extended backward, meet. This represents the apparent position of the camera’s viewpoint.

